# Association of Chronic Acid Suppression and Social Determinants of Health with COVID-19 Infection

**DOI:** 10.1101/2021.01.10.21249545

**Authors:** Bing Zhang, Anna L. Silverman, Saroja Bangaru, Douglas Arneson, Sonya Dasharathy, Nghia Nguyen, Diane Rodden, Jonathan Shih, Atul J. Butte, Wael Noor El-Nachef, Brigid S. Boland, Vivek A. Rudrapatna

**Affiliations:** Division of Gastroenterology and Hepatology, Department of Medicine, University of California, San Francisco, San Francisco, CA 94143; Department of Medicine, University of California, San Diego, La Jolla, CA 92093; Vatche and Tamar Manoukian Division of Digestive Diseases, Department of Medicine, University of California, Los Angeles, Los Angeles, CA 90095; Bakar Computational Health Sciences Institute, University of California, San Francisco, San Francisco, CA 94143; Division of Gastroenterology and Hepatology, Department of Medicine, University of California, San Diego, La Jolla, CA 92093; Department of Neurology, University of California, San Francisco, San Francisco, CA 94143; Center for Data-Driven Insights and Innovation, University of California Health, Oakland, CA

## Abstract

Acid suppressants are a widely-used class of medications previously linked to an increased risk of aerodigestive infections. However, prior studies of these medications as potentially reversible risk factors for COVID-19 have been conflicting. We performed a case-control study involving clinician-abstracted data from 900 health records across 3 US medical centers. We incorporated sociobehavioral predictors of infectious exposure using geomapping to publicly-available data. We found no evidence for an association between chronic acid suppression and incident COVID-19 (adjusted odds ratio 1.04, 95% CI: 0.92-1.17, *P*=0.515). However, we identified several medical and social features as positive (Latinx ethnicity, BMI ≥ 30, dementia, public transportation use, month of the pandemic) and negative (female sex, concurrent solid tumor, alcohol use disorder) predictors of new-onset infection. These results place both medical and social factors on the same scale within the context of the COVID-19 pandemic, and underscore the importance of comprehensive models of disease.

## Introduction

The COVID-19 pandemic, caused by the novel severe acute respiratory syndrome coronavirus 2 (SARS-CoV-2), has affected over 70 million individuals across the globe and killed over 1.6 million as of December 2020, one year after it was first reported in Wuhan, China.^1^ Previous studies have examined risk factors for infection and predictors of disease severity. Established medical risk factors predicting infection and severity encompass age ≥ 65, diabetes mellitus, obesity, smoking, chronic pulmonary disease, cardiovascular disease, chronic kidney disease, malignancy and chronic HIV infection.^2-7^

Proton pump inhibitors (PPIs) and H2-receptor antagonosts (H2RAs) are among the most commonly prescribed medications, used for gastroesophageal reflux disease (GERD) and peptic ulcer disease.^8,9^ Prior work investigating the link between PPIs and aerodigestive infections such as pneumonia has driven the initial hypothesis that PPIs could potentially compromise physiological barriers to SARS-CoV-2 infectivity.^9,10^ Although current medical literature has focused on the identification and quantification of medical-related risk factors, there is a paucity of data on how socioeconomic and behavioral influences on infection risk compared to conventional medical factors. Here, we sought to investigate if there is an association between chronic acid suppression and SARS-CoV-2 infection in patients seen at three urban health systems in California while accounting for concomitant social determinants of health.

## Methods

### Study Design

This is a case-control study involving the review of medical records from three campuses of the University of California (UC) Health system: San Francisco, San Diego, and Los Angeles. The study was approved by the Institutional Review Board (IRB) of the University of California, San Francisco (UCSF; IRB #20-30549). The decision made by UCSF’s IRB was relied upon by the IRB of the University of California, San Diego (UCSD) via a memorandum of understanding (UC IRB Reliance #3499). The decision made by the IRB of the University of California, Los Angeles (UCLA) was that this study did not constitute human subjects research and therefore was exempt.

At the time of the study’s inception in May 2020, deidentified databases of electronic health records (EHR) data at UC Health were queried to to support sample size calculations. At that time, the estimated prevalence of positivity for SARS-CoV-2 virus was 5.6% among those tested across all campuses of UC Health. At UCSF, the prevalence of PPI use was estimated to be 7.5%. We obtained approximate effect sizes from a prior study that assessed the relative risk of acute gastroenteritis among chronic PPI users, motivated in part by work suggesting the possible enteric transmission of the SARS-CoV-2 virus.^11,12^ We calculated that at least 416 cases and controls would be needed to detect two-fold relative risk among chronic PPI users compared to non-users with 80% power. These numbers were rounded up to obtain 450 cases and controls each, or 900 medical records in total. These were divided evenly across all three participating sites (300 records each).

### Patient Selection and Data Collection

Records of all patients who underwent SARS-CoV-2 nasopharyngeal PCR testing at any of the three health systems (UCSF, UCSD, UCLA) from March 1, 2020 to June 10, 2020 were separately retrieved via EHR database query. Stratified uniform sampling was performed to yield 150 cases and 150 controls for manual review at each site. A standardized case report form and accompanying data dictionary were developed. Data elements included PPI and H2RA use, Charlson Comorbidity Index (CCI), additional medical comorbidities and immunocompromised states, and sociodemographic factors (**<u>Supplemental Methods</u>**). Variable definitions and data extraction process was determined and fixed *a priori* and based on consensus among the co-authors.

Medical records of all patients ≥ 18 years old at the time of their initial SARS-COV-2 test were included in the study database. Result of the SARS-CoV-2 PCR testing was determined by documentation in the EHR of each site. For patients who were tested more than once during the data collection period and were consistently negative, the date of the first result was recorded. Patients who tested positive after an initial negative test(s) were recorded as positive, and the date of the first positive test was recorded. Chronic acid suppression was defined as the use of PPI or H2RA daily or as needed for at least four weeks, documented by medical providers in charts or prescription refill records.

All medical history was reviewed and annotated by physicians (BZ, AS, SB, SD, DR). Disagreements during extraction pertaining to the coding of specific variables were adjudicated through discussion and resolved through consensus voting (**<u>Supplemental Methods</u>**). The extraction sheet was modified once to better capture race and ethnicity. Following the completion of data abstraction, datasheets were compiled and subject to quality control measures via both manual (JS) and informatic methods in order to ensure consistent and accurate data capture across all three sites.

### Geomapping Social Determinants of Health

Patient records were mapped to geographic areas using postal zip codes corresponding to their home address. However, most geocoded data including reports from the United States Census utilize zip code tabulation areas (ZCTAs). Postal zip codes were mapped to zip code tabulation areas (ZCTAs), with the method implemented by Williams-Holt.^13-15^

Geocoded covariates related to population and housing density were obtained from the 2010 United States Decennial Census.^16^ Additional geocoded covariates were obtained from the 2018 United States American Community Survey (ACS) using the R package *tidycensus* v0.9.9.2.^17^ For each ZCTA, median household income, public transportation as means of transportation to work for workers 16 and over, and walking as means of transportation to work for workers 16 and over were extracted.

The United States Census Bureau provides Community Resilience Estimates (CREs) at the census tract level defining what percentage of residents in a census tract have 3+ risk factors, 1-2 risk factors, or 0 risk factors.^18^ These risk factors are derived from the 2018 American Community Survey (ACS) and the 2018 National Health Interview Survey (NHIS).

Mask-wearing by county was obtained from a survey conducted by The New York Times.^19^ Briefly, survey responses were collected from 250,000 individuals between 7/2/2020 and 7/14/2020 who were asked: “How often do you wear a mask in public when you expect to be within six feet of another person?” The possible responses were: never, rarely, sometimes, frequently, and always.

Daily social distancing mobility data was obtained from the data company SafeGraph.^20,21^ Briefly, daily anonymized and aggregated mobile device data were obtained from SafeGraph from 2-12-2020 through 7-12-2020. Mobile devices were aggregated by Census Block Groups (CBGs) based on home location and the median percentage of time a device was observed at home versus observed at all during a given time period was calculated for all observed devices in a CBG each day.

SARS-COV-2 Community Mobility Reports were obtained from Google from February 2020 through July 2020.^22^ Briefly, these reports quantify how visits and length of stay at various locations changed compared to a baseline using aggregated and anonymized data from users who opted into Google Location History. Google reported daily values for time spent at the following locations aggregated at the county level: grocery & pharmacy, parks, transit stations, retail & recreation, residential, and workplaces. The time spent at a location was represented as a percent change of time spent at the location compared to the baseline.

The Social Deprivation Index (SDI) has previously been used to quantify levels of disadvantage and evaluate their associations with health outcomes.^23^ The SDI used in this study was developed by the Robert Graham Center and is comprised of seven demographic characteristics from the 2015 ACS.^24^ These characteristics include: income, education, employment, housing, household characteristics, transportation, and demographics. Reported SDI measures for ZCTAs were used in this analysis.

### Outcomes

The. primary outcome was association between chronic acid suppression (PPIs, H2RAs) and risk of SARS-COV-2 infection. Secondary outcomes included the association between SARS-COV-2 infection and the month of testing, demographics, comorbid medical conditions, immunocompromised state, CCI, and social and mobility variables (**<u>Supplemental Table 1</u>**).

### Statistical Analysis

Medical records and covariates with a high degree of missing data (>40%) were excluded from subsequent analysis. Multiple imputation was then performed using the R package *mice v3*.*11*.*0* to impute 10 datasets with 20 iterations per dataset to ensure convergence.^25^

Continuous covariates were converted to binary variables for interpretability in the logistic regression model and corresponding forest plot. To identify binary cut points which may be associated with an increased chance of a positive SARS-COV-2 test, we conducted a grid search of all possible values between the 10^th^ and 90^th^ percentiles for each continuous covariate. The binary cut point associated with the lowest p-value resulting from a chi-square test was used to binarize each continuous covariate.

To identify and select the subset of input covariates that were most relevant to a positive SARS-COV-2 test a multistep process was employed. First, a stepwise linear model to predict a positive SARS-COV-2 test was fit separately for each of the 10 imputed datasets. Both forward and backward selection using the Akaike Information Criterion were used to identify the subset of covariates to be included in the final model for each imputed dataset. Next, covariates which were included in all 10 reduced models were chosen to be included in the final pooled model. Covariates which appeared in ≥50% of the reduced models were considered individually for inclusion in the final pooled model with a Wald test. The selected covariates were used in the final reduced logistic regression model. We initially fit a mixed-effects model on the combined dataset using study site as a random intercept.^26^ These results suggested a low degree of heterogeneity attributable to study site (i^2^ statistic = 0%). Consequently, all subsequent results were obtained by fixed-effect (pooled) models.

### Funding Source

The UCSF Bakar Computational Health Science Institute, the National Institutes of Health, and the Barbara and Joel Marcus UCLA GI Fellowship Research Seed Grant Program had no involvement in the design, collection, analysis, interpretation and decision to publish the current study.

## Results

Of 900 medical records initially selected for manual review, twenty were excluded because they only contained the SARS-COV-2 antibody test (not the nasopharyngeal PCR test), and ten were from patients less than 18 years old at the time of testing **(Figure 1)**. Five individuals with high degrees of missing data (>40%) and four with unknown residential zip codes were excluding due to inability to map geocoded social determinants of health. 861 medical records were retained for downstream analyses.

**Figure 1.**
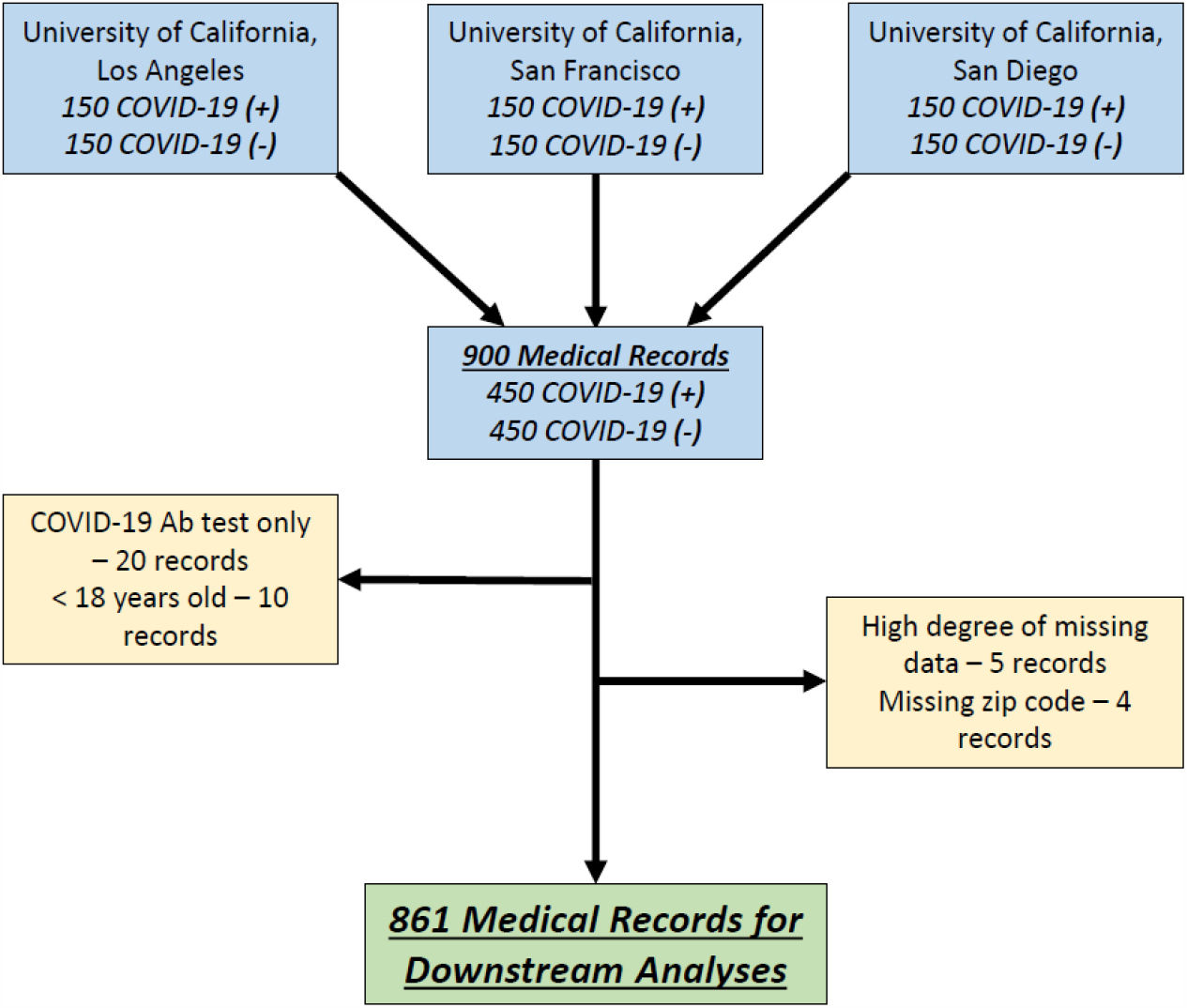
Selection of medical records for study. Selection of medical records was performed independently at three academic hospitals of the University of California. Each site uniformly sampled 150 COVID-19 positive (+) and 150 COVID-19 negative (-) patients. Among these, 861 medial records were retained for downstream analyses.

Distribution of included medical records were similar across the three sites, from which there were 428 (49.7%) documented positive tests **(Table 1)**. Approximately half were female, and one-fifth were ≥65 years old. The majority self-identified as white, and 22% of the total sample identified as Latinx. Most individuals were calculated to have a low CCI (0-2) and were not obese based on body mass index (BMI). Among previously coexisting diseases captured, diabetes (13.7%) and immunocompromised (22.0%) were the most prevalent. Nearly one-fifth tested were healthcare workers, and the majority of people lived in single family homes. During the period sampled, the probability of a positive SARS-CoV2 test decreased significantly after March, likely reflecting the adoption of widespread testing (**Figure 2**).

**Table 1.** Medical and sociodemographic characteristics of medical records included in analysis

| Characteristic | UCLA<br>(N=269) | UCSD<br>(N=297) | UCSF<br>(N=295) | Overall<br>(N=861) |
| --- | --- | --- | --- | --- |
| **Positive SARS-CoV2 test** | 136 (50.6%) | 147 (49.5%) | 145 (49.2%) | 428 (49.7%) |
| **SARS-CoV2 test month** |  |  |  |  |
| Mar | 67 (24.9%) | 35 (11.8%) | 93 (31.5%) | 195 (22.6%) |
| Apr | 103 (38.3%) | 76 (25.6%) | 74 (25.1%) | 253 (29.4%) |
| May | 95 (35.3%) | 146 (49.2%) | 119 (40.3%) | 360 (41.8%) |
| Jun | 4 (1.5%) | 40 (13.5%) | 9 (3.1%) | 53 (6.2%) |
| **Female sex** | 150 (55.8%) | 153 (51.5%) | 161 (54.6%) | 464 (53.9%) |
| **Age - yr** |  |  |  |  |
| >65 yr | 66 (24.5%) | 50 (16.8%) | 56 (19.0%) | 172 (20.0%) |
| ≤65 yr | 203 (75.5%) | 247 (83.2%) | 239 (81.0%) | 689 (80.0%) |
| **Race or ethnic group** |  |  |  |  |
| White | 163 (60.6%) | 212 (71.4%) | 149 (50.5%) | 524 (60.9%) |
| Asian | 22 (8.2%) | 33 (11.1%) | 53 (18.0%) | 108 (12.5%) |
| Black | 25 (9.3%) | 12 (4.0%) | 27 (9.2%) | 64 (7.4%) |
| Other | 21 (7.8%) | 33 (11.1%) | 49 (16.6%) | 103 (12.0%) |
| **Ethnicity Latinx** | 56 (20.8%) | 95 (32.0%) | 38 (12.9%) | 189 (22.0%) |
| **BMI** |  |  |  |  |
| <30 | 170 (63.2%) | 162 (54.5%) | 192 (65.1%) | 524 (60.9%) |
| ≥30 | 64 (23.8%) | 91 (30.6%) | 72 (24.4%) | 227 (26.4%) |
| **PPI use** | 58 (21.6%) | 40 (13.5%) | 33 (11.2%) | 131 (15.2%) |
| **Acid Suppression use** | 66 (24.5%) | 55 (18.5%) | 38 (12.9%) | 159 (18.5%) |
| **Charlson Comorbidity Index** |  |  |  |  |
| 0-2: Mild | 175 (65.1%) | 217 (73.1%) | 221 (74.9%) | 613 (71.2%) |
| 3-4: Moderate | 47 (17.5%) | 36 (12.1%) | 41 (13.9%) | 124 (14.4%) |
| ≥5: Severe | 47 (17.5%) | 44 (14.8%) | 33 (11.2%) | 124 (14.4%) |
| **GERD** | 66 (24.5%) | 43 (14.5%) | 45 (15.3%) | 154 (17.9%) |
| **Previous coexisting disease** |  |  |  |  |
| Heart Failure | 12 (4.5%) | 16 (5.4%) | 10 (3.4%) | 38 (4.4%) |
| Stroke | 14 (5.2%) | 12 (4.0%) | 8 (2.7%) | 34 (3.9%) |
| Dementia | 11 (4.1%) | 10 (3.4%) | 4 (1.4%) | 25 (2.9%) |
| Diabetes | 52 (19.3%) | 32 (10.8%) | 34 (11.5%) | 118 (13.7%) |
| Severe CKD | 17 (6.3%) | 2 (0.7%) | 7 (2.4%) | 26 (3.0%) |
| Solid tumor | 29 (10.8%) | 27 (9.1%) | 21 (7.1%) | 77 (8.9%) |
| AIDS | 2 (0.7%) | 5 (1.7%) | 2 (0.7%) | 9 (1.0%) |
| HIV | 7 (2.6%) | 17 (5.7%) | 14 (4.7%) | 38 (4.4%) |
| Immunocompromised | 78 (29.0%) | 56 (18.9%) | 55 (18.6%) | 189 (22.0%) |
| Autoimmune disease | 11 (4.1%) | 16 (5.4%) | 20 (6.8%) | 47 (5.5%) |
| **Chronic opiate use** | 13 (4.8%) | 30 (10.1%) | 9 (3.1%) | 52 (6.0%) |
| **EtOH use disorder** | 8 (3.0%) | 13 (4.4%) | 11 (3.7%) | 32 (3.7%) |
| **Healthcare worker** | 31 (11.5%) | 64 (21.5%) | 65 (22.0%) | 160 (18.6%) |
| **Residence** |  |  |  |  |
| Single Family Home | 160 (59.5%) | 207 (69.7%) | 173 (58.6%) | 540 (62.7%) |
| Apartment | 91 (33.8%) | 75 (25.3%) | 104 (35.3%) | 270 (31.4%) |
| SNF | 12 (4.5%) | 7 (2.4%) | 6 (2.0%) | 25 (2.9%) |
| Homeless | 2 (0.7%) | 5 (1.7%) | 8 (2.7%) | 15 (1.7%) |
| Cruise | 0 (0%) | 1 (0.3%) | 0 (0%) | 1 (0.1%) |
| **Social Deprivation Index** |  |  |  |  |
| >65 | 128 (47.6%) | 130 (43.8%) | 94 (31.9%) | 352 (40.9%) |
| **Time spent home** |  |  |  |  |
| >85.0% | 250 (92.9%) | 266 (89.6%) | 253 (85.8%) | 769 (89.3%) |
| **Public transportation usage** |  |  |  |  |
| >4.4% | 132 (49.1%) | 106 (35.7%) | 256 (86.8%) | 494 (57.4%) |

**Figure 2.**
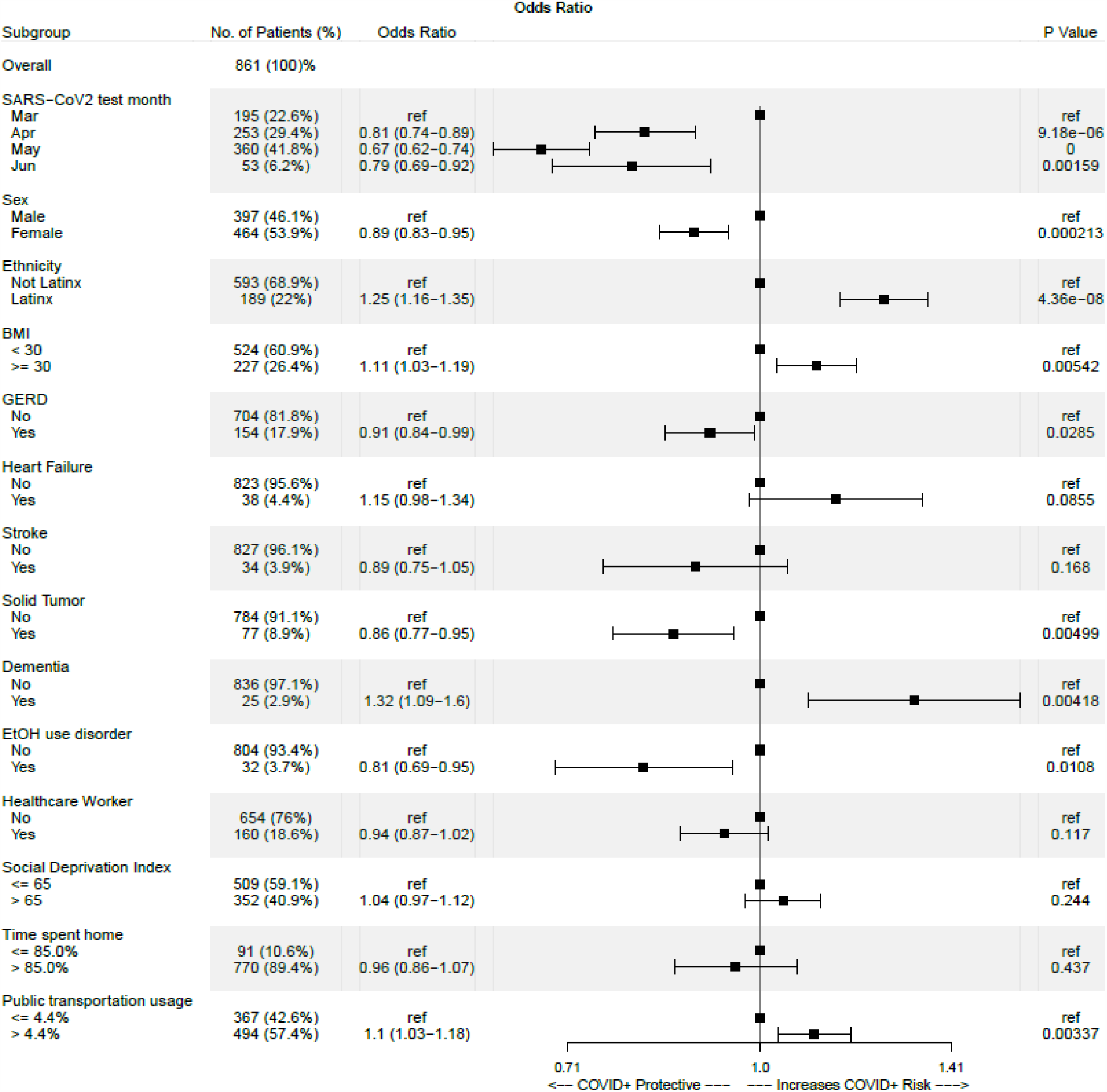
Features identified to correlate with SARS-CoV-2 testing result. Adjusted odds ratios and P-values were calculated when compared to baseline (ref).

### Chronic acid suppression and SARS-CoV-2 risk

To evaluate the effect of chronic acid suppression on SARS-CoV-2 infection risk, logistic model selection using 62 candidate features including chronic PPI use and chronic H2RA use was performed. This process identified neither medication class as significant influences on infection risk (**Figure 2**). This finding was confirmed with manual incorporation of chronic acid suppression use (combination of both PPI and H2RA) into the model (adjusted odds ratio 1.04, 95% CI – 0.92 to 1.17, *P* = 0.515; **<u>Supplemental Figure 1</u>**). Interestingly, GERD was associated with a slightly lower odds of COVID-19 in both models.

### Medical conditions associated with SARS-CoV-2 risk

Several medical comorbidities were identified as significant predictors of incident infection. These included a BMI ≥ 30, and dementia (**Figure 2**). Unexpectedly, non-hematogenous malignancy and history of alcoholic use disorder were associated with a negative test. Other comorbidities including heart failure and stroke were not found to be statistically significant predictors of COVID-19.

### Sociodemographic factors and SARS-CoV-2 risk

Males and individuals of Latinx ethnicity were more likely to have positive test. Finally, those who resided in localities with more than 4.4% of the adult population utilizing public transportation as their main means of commute also were more likely to test positive. Among our sample, healthcare worker status did not strongly correlate with testing outcome, and neither did “social deprivation index” and “time spent at home”.

## Discussion

### Chronic Acid Suppression and SARS-CoV-2 Risk

We identified no association between long-term PPI or H2RA use and COVID-19 after controlling for medical and sociodemographic predictors of infection risk. Our results are consistent with, and complement, two previously published studies utilizing a clinical informatics approach. A case control study using South Korean health-insurance claims reported no assication between current and past use of PPIs and COVID-19.^27^ A cohort study using the United Kingdom’s Biobank also reported no increased risk of COVID-19 among patients who reported taking PPI or H2RA.^28^ Of note, medication data collection period was 2006-2010 for this study, therefore it was not confirmed that participants were still taking these medications following COVID-19 outbreak. By contrast, a single large United States (US) survey-based study found a positive association between adults self-reporting COVID-19 and daily PPI use, but did not find an association with H2RA.^29^ A notable limitation of this study is the reliance of self-reported COVID-19 when testing was available. The study reported a COVID-19 prevalence of 6.4% which was higher than the national estimates at the time, raising concerns regarding the accuracy of self-report. Furthermore, the survey was administered in English, functionally excluding the near quarter of US Census respondents who report limited to no English proficiency.^30^

### Medical Comorbidities and SARS-CoV-2 Risk

Consistent with previous data, our cohort found an increased risk of COVID-19 and a BMI ≥ 30.^31-35^ Furthermore, patients with dementia were more likely to test positive, possibly reflecting frequent interactions with the healthcare sytem or caregivers and a decreased ability to socially distance. Interestingly, those with underlying solid tumors or alcohol use disorder had a lower risk of infection with SARS-CoV-2. We surmised that this may be due to reduced exposure to infected individuals by spending significant time at home and adhering more strictly to precautions, but still having a high need for medical services and therefore receiving more testing. Finally, the negative association between COVID-19 positivity and GERD may reflect functional hearburn, which is comorbid with anxiety and possible proclivity to be tested.^36^

### Sociodemographic Factors and SARS-CoV-2 Risk

An important finding of our study is that members of the Latinx community are at a disproportionate risk of contracting SARS-CoV-2, even after controlling for a variety of medical, socioeconomic and behavioral factors. Current literature in the US and globally has commented mainly on the increased risk of infection, severe disease and death in black patients.^37-44^ Our California-based cohort showed that being of Latinx ethnicity is associated with higher rates of infection with SARS-CoV-2. In addition to data from New York City reporting 34% of COVID-19 deaths in Latinx people despite only representing 29% of the population, our California-based cohort enriched in Latinx patients highlights increased risk.^44,45^ In the 45 states reporting data by ethnic group, 20 report the proportion of cases among Latinx people is twice as high and 11 report it is three times as high as would be expected based on the general population.^46,47^ These collective data underscore the substantial vulnerability of this ethnic group.

Patient zip codes and geolocation data provided indirect insight into the physical environments and behaviors of the communities in which the patients reside and evaluated effects of social determinants of health on COVID-19 risk. We found patients living in zip codes with increased public transportation reliance were associated with SARS-COV-2 positivity. Since the ability to work from home, avoid public transportation, financially accommodate for furloughs from work are not equally distributed, the ability to social distance effectively may be one of privilege.^44,47,48^ Although these differences in social determinants of health are receiving specific attention in the current pandemic, they speak to important and systematic disparities across healthcare that need to be better understood and addressed.

In our cohort, females had a significantly lower probability of infection with SARS-CoV-2. It is currently unknown if there are immunologic or behavioral explanations why female gender may be protective.^49^ Future work is needed to clarify this finding.

### Strengths and Limitations

Our study has several strengths. The multi-center design incorporates patients from three large urban academic health systems spanning Northern and Southern California providing care for a diverse patient cohort. Data included confirmed SARS-COV-2 by PCR test results in addition to medications and comorbidities data collected and curated by physicians to improve accuracy. Application of geolocation using zipcode enabled identification of sociodemographic determinants of infection risk. An important limitation of our study is possible selection bias. Our study cohort consisted only of tested patients. Attempts to correct this include incorporating multiple predictors of the probability of testing as well as the probability of infection itself; nonetheless we cannot exclude residual bias. Other potential limitations include possible measurement bias (e.g. discordance between EHR documentation and actual patient use of medications), ecological fallacy (i.e. attribution of locality-level data to individuals), and cohort unrepresentativeness (i.e. differences between urban, tertiary-care cohorts and the general US population). It should be noted that testing was performed both for symptom-triggered testing and preprocedural or hospitalization screening. Thus, the pretest probability was not homogeneous. Associations identified in this observational study should not be assumed causal and require prospective validation.

## Conclusion

This multicenter case-control study demonstrated no association between chronic acid suppression use and risk of COVID-19. In addition, we found that Latinx ethnicity and residing in a zip code with high public transportation usage were associated with increased risk of COVID-19 which is in keeping with growing evidence that this pandemic has disproportionately affected vulnerable communities in our society. Further work is needed to address reducing the burden of the SARS-COV-2 pandemic on these communities.

## Data Availability

The data used for this study contains protected health information, including dates and zip codes. As such, the underlying data have not been made available for reuse. However, the analytic codes along with randomly generated demo data and detailed tutorial have been made publicly available on github. Datasets supporting analytical workflow of this manuscript is also available.

https://github.com/darneson/CovidPPI

https://figshare.com/articles/dataset/Datasets_supporting_analytical_workflow_of_Chronic_Acid_Suppression_and_Social_Determinants_of_COVID-19_Infection/13380356

## Acknowledgments

The authors acknowledge the US Census, The New York Times, SafeGraph, and Google LLC for providing data relevant to this study. They thank the leadership and governance of the UC Office of the President and UC Health, as well as the Clinical and Translational Science Institutes of all member campuses for their support of the clinical informatics infrastructure necessary for this work.

## Data Availability

The data used for this study contains protected health information, including dates and zip codes. As such, the underlying data have not been made available for reuse. However, the analytic codes along with randomly generated demo data and detailed tutorial have been made publicly available on github at: https://github.com/darneson/CovidPPI. Datasets supporting analytical workflow of this manuscript is available at: https://figshare.com/articles/dataset/Datasets_supporting_analytical_workflow_of_Chronic_Acid_Suppression_and_Social_Determinants_of_COVID-19_Infection/13380356.

## Competing Interest Statement

*The authors declare no competing interests*. BSB has received consulting fees from Bristol Myers Squibb, Pfizer, and Takeda, unrelated to current research. VAR has received support from Janssen Inc. for research unrelated this work. AJB is a co-founder and consultant to Personalis and NuMedii; consultant to Samsung, Mango Tree Corporation, and in the recent past, 10x Genomics, Helix, Pathway Genomics, and Verinata (Illumina); has served on paid advisory panels or boards for Geisinger Health, Regenstrief Institute, Gerson Lehman Group, AlphaSights, Covance, Novartis, Genentech, and Merck, and Roche; is a shareholder in Personalis and NuMedii; is a minor shareholder in Apple, Facebook, Alphabet (Google), Microsoft, Amazon, Snap, 10x Genomics, Illumina, CVS, Nuna Health, Assay Depot, Vet24seven, Regeneron, Sanofi, Royalty Pharma, AstraZeneca, Moderna, Biogen, Paraxel, and Sutro, and several other non-health related companies and mutual funds; and has received honoraria and travel reimbursement for invited talks from Johnson and Johnson, Roche, Genentech, Pfizer, Merck, Lilly, Takeda, Varian, Mars, Siemens, Optum, Abbott, Celgene, AstraZeneca, AbbVie, Westat, and many academic institutions, medical or disease specific foundations and associations, and health systems. AJB receives royalty payments through Stanford University, for several patents and other disclosures licensed to NuMedii and Personalis. AJB’s research has been funded by NIH, Northrup Grumman (as the prime on an NIH contract), Genentech, Johnson and Johnson, FDA, Robert Wood Johnson Foundation, Leon Lowenstein Foundation, Intervalien Foundation, Priscilla Chan and Mark Zuckerberg, the Barbara and Gerson Bakar Foundation, and in the recent past, the March of Dimes, Juvenile Diabetes Research Foundation, California Governor’s Office of Planning and Research, California Institute for Regenerative Medicine, L’Oreal, and Progenity. All other authors have nothing to disclose relevant to the manuscript. The disclosures had no involvement in the design, collection, analysis, interpretation and decision to publish.

## Funding Source

This study was supported by funding from the UCSF Bakar Computational Health Science Institute and the National Center for Advancing Traanslational Sciences of the NIH, grant number UL1TR001872 (UCSF), UL1TR001881 (UCLA), and UL1TR001442 (UCSD). UCLA’s effort was supported by the Barbara and Joel Marcus UCLA GI Fellowship Research Seed Grant Program. BZ is supported by NIH/NIDDK, grant number T32DK007007-45. NN is supported by NIH/NIDDK, grant number T32DK007202. BSB is supported by NIH/NIDDK, grant number K23DK123406. VAR was supported by NIH/National Center for Advancing Translational Sciences, grant number TL1TR001871.

## Notes

### Author Declarations

UCSF IRB, UCLA IRB, UCSD IRB

