## Supplemental Data for "Association of Chronic Acid Suppression and Social Determinants of Health with COVID-19 Infection"

**Supplemental Figure 1** – Inclusion of chronic acid suppression use to features identified to correlate with SARS-CoV-2 testing results

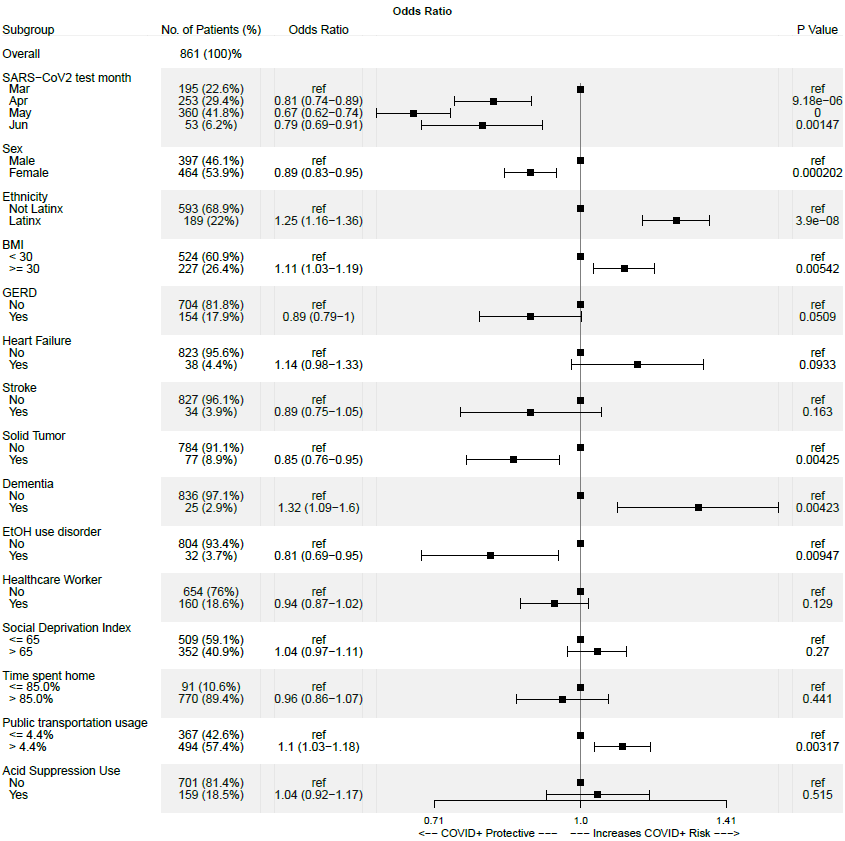

Adjusted odds ratios and P-values were calculated when compared to baseline (ref).

**Supplemental Table 1 –** Features included in logistics regression model

Location (Site)

Date of COVID-19 Test

Age

Sex

Race

Ethnicity

PPI Use

H2RA Use

GERD/Reflux

Myocardial Infarction

Congestive Heart Failure

Peripheral Vascular Disease

Stroke

Dementia

COPD

Connective Tissue Disease

Peptic Ulcer Disease

Liver Disease

Diabetes Mellitus

Hemiplegia

Moderate to Severe CKD

Solid Tumor

Leukemia

Lymphoma

AIDS

Hypertension

Asthma

HIV

CKD (Cre < 3)

BMI

IBD

Autoimmune

Hx Cancer

Chronic opiate use

Immunocompromised (by med or comorbidity)

Smoker (including marijuana)

EtOH use disorder

Healthcare worker

Apartment (living situation)

CCI

Acid Suppression Use

Median Income (Zip code - US Census)

Public Transportation (Zip code - US Census)

Walk to Work (Zip code - US Census)

Population Density (Zip code - US Census)

Housing Density (Zip code - US Census)

Percent Low Risk Factors (Zip code - Community Resilience Estimates)

Percent Moderate Risk Factors (Zip code - Community Resilience Estimates)

Percent High Risk Factors (Zip code - Community Resilience Estimates)

Mask Use: Never (County - New York Times)

Mask Use: Rarely or More Often (County - New York Times)

Mask Use: Sometimes or More Often (County - New York Times)

Mask Use: Frequently or More Often (County - New York Times)

Mask Use: Always (County - New York Times)

Time Spent At Retail Stores (County - Google; as a percentage of baseline)

Time Spent At Grocery Stores (County - Google; as a percentage of baseline)

Time Spent At Parks (County - Google; as a percentage of baseline)

Time Spent At Public Transit (County - Google; as a percentage of baseline)

Time Spent At Workplaces (County - Google; as a percentage of baseline)

Time Spent At Residential Areas (County - Google; as a percentage of baseline)

Social Deprivation Index (Zip code)

Median Percentage of Time Spent Home (Zip code - SafeGraph)

**Supplemental Methods –** Chart Extraction Variables

1. Location of test: 0 = UCSF, 1 = UCSD, 2 = UCLA
2. Patient MRN: enter MRN (note that this column exists only for internal use and can be deleted prior to transfer of data to UCSF or other site)
3. COVID-19 DX: 0 = no; 1 = yes
4. Date of COVID-19 test: mm/dd/yyyy
5. Age: enter age at the time of the COVID-19 test
6. Sex: 0 = male, 1 = female. If neither of the above leave blank
7. Race: 0 = White, 1 = Black or African American, 2 = American Indian or Alaska Native, 3 = Asian, 4 = Native Hawaiian or Other Pacific Islander
8. Ethnicity: 0 = not Hispanic or Latino; 1 = Hispanic or Latino
9. Zip code: enter zip code
10. Chronic PPI or H2 Blocker (defined as more than 4 weeks): 0 = no; 1 = yes
11. PPI Type: enter the generic name of the PPI that the patient is on at the time of COVID test, or NA if unknown
12. PPI Blocker Dose (mg): enter dose or NA if unknown
13. PPI Frequency: 0 = once daily; 1 = twice daily, NA = unknown
14. Duration of PPI Use (months): enter the total duration of time (in months) with regular use of PPI. If the patient is a PRN user of PPI, annotate as **-1**. If the number of months is unknown but the number of years is documented (e.g. 2 years), report as the equivalent number of months (e.g. 24 months). If the start date of PPI is unclear by chart review annotate as NA
15. H2 Blocker: 0 = no, 1 = yes
16. H2 Blocker Type: type name of H2 blocker that the patient is on at the time of COVID test
17. H2 Blocker Dose (mg): enter number
18. H2 Blocker Frequency: 0 = once daily; 1 = twice daily
19. Duration of H2 Blocker use (months): enter number
20. GERD/Reflux/Regurgitation: 0 = no; 1 = yes

**CCI**

21. MI (anytime in the past prior to COVID Test): 0 = no; 1 = yes. CAD in the absence of myocardial death (defined by EKG, Echocardiography, PET, MRI or equivalent) should NOT be treated as evidence of an MI. Impaired myocardial function (e.g. wall motion abnormalities) may be treated as a ‘yes’ for this category even if reversible ischemia has not been clearly established.
22. CHF (anytime in the past prior to COVID Test): 0 = no; 1 = yes. This may include conditions with structural heart damage commonly characterized by decreased ejection fraction but may also include conditions with persevered ejection fraction (i.e. ‘diastolic heart failure’). Many cardiomyopathies will fall under this category.
23. Peripheral Vascular Disease (anytime in the past prior to COVID Test): 0 = no; 1 = yes. These include peripheral arterial disease and as well as venous insufficiency.
24. Stroke (CVA or TIA) (anytime in the past prior to COVID Test): 0 = no; 1 = yes. These encompass both ischemic and hemorrhagic varieties, and overall reflect an increased risk of pneumonia due to their association with dysphagia and aspiration.
25. Dementia (anytime in the past prior to COVID Test): 0 = no; 1 = yes
26. COPD (anytime in the past prior to COVID Test): 0 = no; 1 = yes
27. Connective Tissue Disease (anytime in the past prior to COVID Test): 0 = no; 1 = yes. These generally refer to autoimmune conditions of the skin, joints, and soft tissue but may encompass other entities. If edge cases arise they can be discussed and adjudicated in group meetings.
28. Peptic Ulcer Disease: 0 = no; 1 = yes. Patients for whom these ulcers have resolved (e.g. NSAID discontinuation, H Pylori treatment with confirmation of clearance) can be treated as ‘no’.
29. Liver Disease: 0 = no; 1 = yes. Self-limited or resolved liver diseases (e.g. Hep A, Drug-induced liver injury) should not be treated as ongoing liver disease.
30. DM (1 or 2): 0 = none or diet-controlled, 1 = uncomplicated, 2 = end-organ damage
31. Hemiplegia (anytime in the past prior to COVID Test): 0 = no; 1 = yes
32. Creatinine ≥3, dialysis, s/p kidney transplant, uremia (most recent lab value prior to COVID test): 0 = no; 1 = yes
33. Solid Tumor: 0 = none, 1 = localized, 2 = metastatic
34. Leukemia: 0 = no; 1 = yes
35. Lymphoma: 0 = no; 1 = yes
36. AIDS: 0 = no; 1 = yes, NA if unknown. Annotate based on the most recent CD4 count of the patient if available within the 1y prior to a COVID-19 test, with AIDS defined as the presence of HIV positivity (by PCR or serologies) with an absolute CD4 count less than 200.
37. CCI Score: calculated from above using - <https://www.mdcalc.com/charlson-comorbidity-index-cci>

**Other Comorbidities**

38. HTN (anytime in the past prior to COVID Test): 0 = no; 1 = yes. HTN should be clearly noted by a primary care note. At the reviewer’s discretion, this diagnosis may be ascertained in the setting of repeated elevations in ambulatory blood pressure (>140/90). Missing data should be rare for this category.
39. Asthma: 0 = no, 1 = yes.
40. HIV: 0 = no, 1 = HIV, 2 = AIDS.
41. CKD (Cre < 3): 0 = no, 1 = yes.
42. Medications: immunocompromised medications.
43. BMI: enter number. If BMI as captured within the last year prior to COVID19 testing is not explicitly reported in the EHR, but weight is, use the following website. Treat height as a fixed variable (ok to use old height measurements) https://www.nhlbi.nih.gov/health/educational/lose_wt/BMI/bmicalc.htm
44. IBD (anytime in the past prior to COVID Test): 0 = no; 1 = yes
45. Autoimmune disease (anytime in the past prior to COVID Test): 0 = no; 1 = yes
46. Hx Cancer (anytime in the past prior to COVID Test): 0 = no; 1 = yes. These typically will encompass both solid and hematological malignancies. Patients who have had cutaneous malignancies (e.g. basal cell cancer) which have been excised and deemed cured may be treated as a 0. Other malignancies that were identified and definitively resected at an early stage without need for systemic chemotherapy may be discussed in group with the possibility of revising this variable: it is intended to capture states of immunocompromise related to history of cancer treatment.
47. Type of cancer: free text.
48. Prior cancer Tx: 1 = chemo (including immunotherapy); 2 = bone marrow transplant, 3 = other
49. Chronic Opiate Use: 0 = no; 1 = yes
50. Immunocompromised (by med or comorbidity): 0 = no, 1 = yes
51. Recent antibiotic use (< 3mos): 0 = no; 1 = yes
52. Smoker (including marijuana): 0 = none, 1 = prior, 2 = current, NA if missing
53. EtOH use disorder: 0 = none, 1 = prior, 2 = current, NA if missing
54. Healthcare worker: 0 = no, 1 = yes
55. Group Living (SNF, rehab, shelter, etc): 0 = no; 1 = yes, NA if missing
56. Apartment: 0 = no; 1 = yes

**Supplemental Methods –** Discussion on Coding of Specific Variables

1. Autoimmune diseases – IBD, ankylosing spondylitis, myasthenia gravis, type 1 diabetes, rheumatoid arthritis; hypothyroidism is not charted as autoimmune
2. Every other day PPI – code as PRN
3. Immunosuppression medications – steroids, biologics, small molecules, chemotherapy, MTX/6MP
4. Diabetes and cirrhosis should be considered as immunocompromised
5. For CCI – current solid tumors are recorded as yes
6. Basal cell cancer – not counted as solid cancer
7. The most recent PPI should be recorded on datasheet
8. MI – counted if there is history of NSTEMI or STEMI
